# Physiological (TCR-like) regulated lentiviral vectors for the generation of improved CAR-T cells

**DOI:** 10.1101/2021.03.17.21253300

**Authors:** María Tristán-Manzano, Noelia Maldonado-Pérez, Pedro Justicia-Lirio, Pilar Muñoz, Marina Cortijo-Gutiérrez, Kristina Pavlovic, Rosario Jiménez-Moreno, Sonia Nogueras, MDolores Carmona, Sabina Sánchez-Hernández, Araceli Aguilar-González, María Castella, Manel Juan, Concepción Marañón, Karim Benabdellah, Concha Herrera, Francisco Martin

## Abstract

**Background:** Chimeric antigen receptor (CAR) T cells directed against CD19 have achieved impressive outcomes for the treatment of relapsed/refractory B lineage lymphoid neoplasms. However, CAR-T therapy still has important limitations due to severe side effects and the lack of efficiency in 40-50% of the patients. Most CARs-T products are generated using retroviral vectors with strong promoters. However, high CAR expression levels can lead to tonic signalling, premature exhaustion and over-stimulation of CAR-T cells, reducing efficacy and increasing side effects. TCR-like expression of the CAR through genome editing resulted in enhanced anti-tumour potency, reducing tonic signalling and improving CAR-T phenotype. In this manuscript, we searched for LVs that mimic the TCR expression pattern as a closer-to-clinic alternative for the generation of improved CAR-T cells.

**Methods:** Different LVs containing viral and human promoters were analysed to select those that closely mimic a TCR-like kinetic profile upon T-cell activation. *WAS* gene proximal promoter-driven LVs (AW-LVs) were selected to express a second generation 4-1BB aCD19 CAR (ARI-0001) into T cells to generate AWARI CAR-T cells. TCR-like AWARI and EF1α-driven ARI CAR T cells were analysed for *in vitro* and *in vivo* killing efficiency using leukaemia and lymphoma cellular models. Tonic signalling, exhaustion markers and phenotype were determined by flow cytometry. Large-scale automated manufacturing of AWARI CAR-T cells was performed in a CliniMACs Prodigy bioreactor.

**Results:** Our data showed that LVs expressing the transgene through the *WAS* gene proximal promoter mimic very closely the TCR (CD3) expression pattern kinetic upon TCR stimulation or antigen encounter. Compared to EF1α-driven ARI CAR-T cells, AWARI CAR-T cells exhibited a higher proportion of naïve/stem cell memory T cells with less exhausted phenotype after efficient killing of CD19+ cells both *in vitro* and *in vivo*. AWARI CAR-T cells also showed lower tonic signalling and reduced secretion of pro-inflammatory cytokines and were efficiently manufactured in large-scale GMP-like conditions.

**Conclusions:** *WAS-gene*-promoter driven LVs can be used to generate physiological 4-1BB-CAR-T cell products with lower tonic signalling, improved phenotype and a safer profile. We propose the use of TCR-like LVs as an alternative to strong-promoter driven LVs for the generation of CAR-T products.

## BACKGROUND

Chimeric antigen receptor (CAR) T cells have become one of the most promising approaches for the treatment of cancer in particular for B lineage lymphoid neoplasms using αCD19-CAR-T cells, as emphasized with four FDA/EMA-approved Advance Therapy Medicinal Products (ATMPs): tisagenlecleucel (Kymriah, Novartis), axicabtageneciloleucel (Yescarta, Kite-Gilead), brexucabtagene autoleucel (Tecartus, Kite-Gilead) and lisocabtagenemaraleucel (Breyanzi, Bristol Myers Squibb). However, in spite of the impressive results of αCD19-CAR-T cells, there are still several aspects that must be improved. Aggressive severe side effects due to CAR-T overstimulation, such as cytokine release syndrome (CRS) and neuroinflammation are common among treated patients and can lead to fatal outcomes. Furthermore, sustained complete remissions range from 62% to 42% for patients treated with commercial ATMPs, evidencing much room for improvement^1^.

Different works have uncovered the importance of controlling CAR expression levels at the surface of the CAR-T cells in order to optimize its therapeutic activity ^2-4^. In spite of this, all four approved ATMPs approved and most of those that are been tested in ongoing clinical trials use autologous T cells transduced with retroviral vectors expressing a αCD19-CAR through constitutive, strong promoters such as the human EF1α and the murine stem cell virus LTR (MSCV). High CARs concentrations on the T cell surface can result in spontaneous clustering of the CARs (independent of the ligand) leading to tonic-signalling^45^. This tonic signalling can affect safety (pro-inflammatory cytokines secretion in non-target tissues) and efficacy (premature exhaustion due to continuous proliferation) of CAR-T cells^3-5^. Furthermore, high-density and constitutive CAR presence on the T cell surface can also lead to overstimulation upon antigen recognition that also affect safety (excess of pro-inflammatory cytokine secretion) and efficacy (early exhaustion, apoptosis and loss of T_N/SCM_ phenotype) of the CAR-T cells^2, 3^. In this direction, Eyquem et al. generated CAR-T cells expressing the αCD19-28ζ CAR through the endogenous T cell receptor constant alpha chain (TRAC) locus using CRISPR/Cas9^2^. In this manuscript, the authors concluded that tight transcriptional regulation of CAR expression, lowering CAR levels upon target binding and recovering 1-3 days later, was critical for optimal CAR-T performance. However, despite the great potential of genome editing tools for therapeutic applications, there are still several technological and safety issues that need to be solved before the approval of genome edited cells as ATMPs.

Contrary to genome editing tools, lentiviral vectors (LVs) derived from HIV-1 have already been approved by the FDA and EMA (Kymriah, Breyanzi and Zynteglo). Latest generation LVs are very resistant to transgene silencing^6,7^, and allow the control of the transgene through physiological or drug inducible promoters^8-13^. We therefore searched for LVs that express transgenes on T cells following a TCR expression kinetic. Since the TRAC promoter in human mature T cells is not well defined, we focus in a well-defined promoter that controls the expression of the WASP protein, which is involved in the formation of the immunological synapse and in translating TCR signals to several T cells functions^14,15^. We reason that *WAS-* promoter driven LVs ^8 11 16^ could be an interesting option to express CARs due to their moderate expression levels and the functional relationship with the TCR. Here we showed that, indeed, *WAS*-promoter driven LVs partially mimicked the TCR expression kinetic both in eGFP- and CAR-expressing LVs, with a small down-regulation upon stimulation and recovering basal levels in 5-7 days. Based on these data, we generated TCR-like CAR-T cells using LVs and analysed potential improvements compared to standard CAR-T cells expressing the CAR through strong promoters. TCR-like expression of a αCD19-CAR lead to lower tonic signalling in the absence of antigen as well as a better response in its presence, with lower exhaustion markers, lower proinflammatory cytokine secretion and improved phenotype both *in vitro* and *in vivo*.

## MATERIALS and METHODS

### Cell culture

Nalm6 (ATCC® CRL-3273), Namalwa (ATCC® CRL-1432), Jurkat (ATCC® TIB-152), HL-60 (ATCC® CCL-240) and HEK-293T (ATCC® CRL-11268) cells were cultured as described elsewhere. Namalwa and Nalm6 were modified to express enhanced GFP (eGFP) and Nanoluciferase (NanoLuc) using the SELWP, as described previously^17^.

### Human samples

Primary T cells were isolated from fresh or frozen apheresis products obtained from healthy donors at the Hematology Department of the Hospital Universitario Reina Sofía (Córdoba, Spain). All donors gave their written informed consent and the study was performed according to the guidelines of the local ethics committee and complies with the requirements regarding quality and safety for donation, obtaining, storage, distribution, and preservation of human cells and tissues under the Spanish specific regulation (RD-L 9/2014). Pan-T cells were isolated by negative selection using immunomagnetic beads (Pan T cell Isolation Kit, Miltenyi Biotec) and following MACSExpress Separator (Miltenyi Biotec) or AutoMACs Pro Separator’s (Miltenyi Biotec) protocol and cultured in TexMACS (Miltenyi Biotec) supplemented with 20 UI/ml of IL-2 (Miltenyi Biotec). T cells were activated with T Cell TransAct (Miltenyi Biotec).

### Plasmid constructs

To construct the SELWP vector, a self-inactivated (SIN) LV expressing eGFP and NanoLuc under the spleen focus-forming virus (SFFV) promoter, an eGFP-P2A-NanoLuc (NanoLuc sequence obtained from GeneBank accession: JQ437370. Nucleotides 100-616) flanked by AscI/SbfI restriction sites was designed and synthesized by Genscript (Genscript USA Inc. NY, U.S.A). The eGFP-P2A-NLuc was cloned into the SEWP LV ^18^ by standard molecular biology techniques to obtain the SELWP.

We used self-inactivated LVs-driven eGFP already available in our laboratory. SE^16^, CEWP^10^ and EFEWP (our laboratory) driving eGFP under the control of SFFV, cytomegalovirus, and EF1a promoters, respectively. *WAS*-promoter based LVs: WE-LVs harbours 0.5kb of the *WAS* proximal promoter^16^, and the AWE-LVs include 0.5kb of the *WAS* proximal promoter and 0.38 kb of the alternative promoter^8^.

For AWARI LVs generation, the AW promoter were obtained from the AWE ^8^ by ClaI / BamHI and inserted into the ARI-0001 plasmid^19^, replacing the EF1a promoter.

### Lentiviral vector production and titration

Lentiviral vectors were generated by co-transfection of HEK-293T cells with the plasmid of interest, the plasmid pCMVDR8.91and the p-MD-G plasmid as previously described^12^. LVs were concentrated by ultracentrifugation at 90.000g for 2h at 4°C, resuspended in TexMACs and storaged at −80°C. LVs titters were determined by transducing Jurkat cells with different dilutions of viral supernatant. Percentage of positive cells was determined by flow cytometry and transducing units per ml (TU/ml) were estimated according to the formula: (10^5^ plated cells × % of positive cells)]/ml of LV.

### T cell transduction

Activated primary T cells were transduced with the different LVs at a multiplicity of infection (MOI) of 10 through spinoculation (800 x g for 60 min at 32°C). Media was exchanged after 5 hours of incubation. At 4-6 days after transduction, the percentage of transduced cells was determined by flow cytometry.

### Expression pattern analysis of LVs on T cells

For T cells transduced with eGFP-LVs, 10^5^ T cells were stimulated with TransAct (Miltenyi Biotec) after 10 days of the initial activation for transduction. Similarly, 10^4^ transduced CAR-T cells were stimulated with 10^4^ CD19+ Namalwa-GFP-NLuc cells in order to simulate the CAR/TCR signalling axis in U-bottom 96-wells plates. eGFP or CAR expression was determined at different time-points after stimulation. Cells were stained and fixed with 2% PFA prior FACS acquisition. Expression was indicated as the ratio of MeFI of positive population against the MeFI of non-transduced total population. Fold change indicates the ratio of expression or percentage related to those values at 0h.

### Flow cytometry

CAR expression was determined with a biotin-conjugated goat anti-murine Fab SP-longer Spacer IgG (Jackson Immunoresearch) and APC-conjugated streptavidin (ThermoFisher). For human T cell phenotyping the following mAbs were used: CD45RA-PE/FITC (HI-100), CD62L-PE-Cy7 (DREG56), CD3-PerCP-Cy5/APC-780 (OKT3), PD1-APC (MIH4), LAG-3-PE (3DS223H), TIM-3-APC-Cy7 (F38-2E2) all from eBioscience (ThermoFisher). Tonic signalling was determined by intracellular staining with pCD3z-PE (Tyr142, 3ZBR4S) and Fix & Perm Kit (Nordic Bio). Samples were acquired on a FACSCantoII cytometer. FlowJo software (TreeStar) was used for data analysis.

### Cytokine secretion of CAR-T cells

To analyse cytokine production after T cells stimulation, 5×10^4^ CAR-T cells were co-cultured with Namalwa-GFP target cells at ratio 1:1 in TexMACs without any supplement. Supernatants were collected after 24h and frozen at −80°C. TNFα and IFN□ were measured with anti-human TFN-α Ready-SET-Go! Kit of Affymetrix (eBioscience) and ELISA MAX Deluxe Set (Biolegend) respectively following manufacturer’s instructions.

### Cytotoxicity assay

Target cells (CD19+), such as Namalwa and Nalm-6 cells expressing GFP-Nluc, and non-target cells (CD19-) HL60 cells (unlabelled), were co-cultured at a concentration of 5×10^3^ cells per well in duplicate in 96-well U-bottom plates, and incubated with T cells at various effector target (E:T) ratios (0.5:1, 1:1, 2:1) in non-supplemented TexMACs during 16h or 48h as indicated. Percentage of lysis was determined by flow cytometry related to basal lysis produced by non-transduced T cells. Specific lysis was determined as follows: 1-(%CD19+ /% HL-60 in CAR-T cells condition/ %CD19+/%HL-60 in NTD condition) x100.

### *In vivo* xenograft animal model and bioluminescence analysis

10 to 12-weeks-old NOD/scid-IL-2Rnull (NSG) mice were inoculated intravenously with 3×10^5^ Namalwa-GFP-NLuc cells per mice, and three days later, mice were randomly infused with CAR-T cells (5×10^6^ or 10×10^6^), non-transduced T cells (5×10^6^ or 10×10^6^) or PBS in the tail’s vein. Re-challenge was assessed by re-inoculating intravenously a new dose of Namalwa-GFP-Nluc cells. Our experiments are based on a modified randomized block design in which each block receives more than one treatment at different periods. Subjects were randomly assigned to receive the different treatments (PBS, T cells or CAR-T cells). This will allow a comparison between the different treatment groups in pairs (each one compared to the control and each one compared to each other). For the estimation of the sample size in each experiment, we calculated the sample size necessary to obtain a p ≥ 0.05 to be of n ≥ 5 for controls/CAR-T comparison and an n ≥ 8 for AWARI/ARI comparisons based on Mayer et al^20^. For bioluminescence analysis, furimazine (Nano-Glo, Promega) was diluted at 1/60 in PBS and injected intraperitoneally immediately prior to acquisition on an IVIS Spectrum analyzer (Caliper, Perkin Elmer). Images were acquired during 180s, open field, and analysed using the Living Image 3.2 (Perkin Elmer) or AURA Imaging Software 3.2 (Spectral Instruments Imaging). Mice were sacrificed if experiencing a weight loss greater than 20% or at the indicated days. Samples of blood, bone marrow, liver, spleen and brain were collected. Blood was extracted and diluted 1/5 in EDTA. Cells were obtained from liver and spleen by mechanical disruption and from bone marrow by the perfusion of both femurs and tibias. Brain’s cells were obtained after Percoll-gradient separation. Percentage of surviving Namalwa-GFP-Nluc and T cells (hCD3+) were determined by FACS in the singlet’s gate and then in “human cells” gate, which was previously established after acquiring an artificial mixture of Namalwa cells and human T cells used as control.

### GMP-like manufacturing of CAR-T cells on CliniMACs Prodigy

Large scale manufacturing of CAR-T cells on CliniMACs Prodigy was carried out under GMP-like conditions into Gene-Cell Therapy cleanrooms of Cell Therapy Unit of Hospital Universitario Reina Sofía (Córdoba, Spain). Two different apheresis from a healthy donor was thawed and around 100×10^6^ T cells were inoculated into the CliniMACs prodigy bioreactor (Miltenyi Biotec). CD4 and CD8 cells were selected with CD8 and CD4 Reagent (Miltenyi Biotec), cultured with IL-7/IL-15 (Miltenyi Biotec) and activated with αCD3/αCD28 GMP T cell TransAct (Miltenyi Biotec). At day 2 of the process, these cells were transduced with AWARI-LVs (MOI=5). Cells were cultured in TexMACs GMP medium containing GMP-grade IL-15 and IL-7 (Miltenyi Biotec) for 9 or 10 days. Final product was collected with 100ml of NaCl 0,9% + 0,5% human serum albumin (HSA).

Cells were stained with CD3-APC, CD4-FITC, CD8-APC Vio770, CD14-PE Vio770, CD45-Vioblue (Miltenyi Biotec). To assess the efficiency of transduction CAR-T cells were stained with CD-19 Biotin and Anti-biotin PE (Miltenyi Biotec). Viability was tested by using 7AAD (Miltenyi Biotec). Phenotype was determined with CD45RA-APC (HI100) and CCR7-BV421 (2-L1-A RUO) from BD Pharmingen. Cells were acquired on a MACsQuant cytometer (Miltenyi Biotec) and analyzed with MACsQuantify Analysis Software (Miltenyi Biotec).

### Data analysis

Statistical analyses were performed using GraphPad 1.6 software (GraphPad Software Inc., La Jolla, CA). Data were expressed as the mean ± SEM. Each performed statistical test was indicated in every figure caption. Survival curves were constructed using the Kaplan-Meier method.

## RESULTS

### *WAS* promoter-driven LVs mimic the TCR expression pattern upon T-cell activation

As LVs have already been approved by EMA and the FDA, LVs expressing the CARs with a similar kinetic than the TCR (TCR-like LVs) could be a real alternative to existing LVs to generate more potent and safer CAR-T cells. We first investigated whether *WAS*-promoter-driven LVs could have a more physiological expression pattern compared to current LVs used for CAR-T cells generation. We generated eGFP-expressing LVs particles from two different *WAS*-pronioter-driven LVs (**Fig. 1A**, WE and AWE)^8, 16^ and three LVs driven by strong promoters (**Fig. 1A**, CEWP, SE and EFEWP).

**Figure 1.**
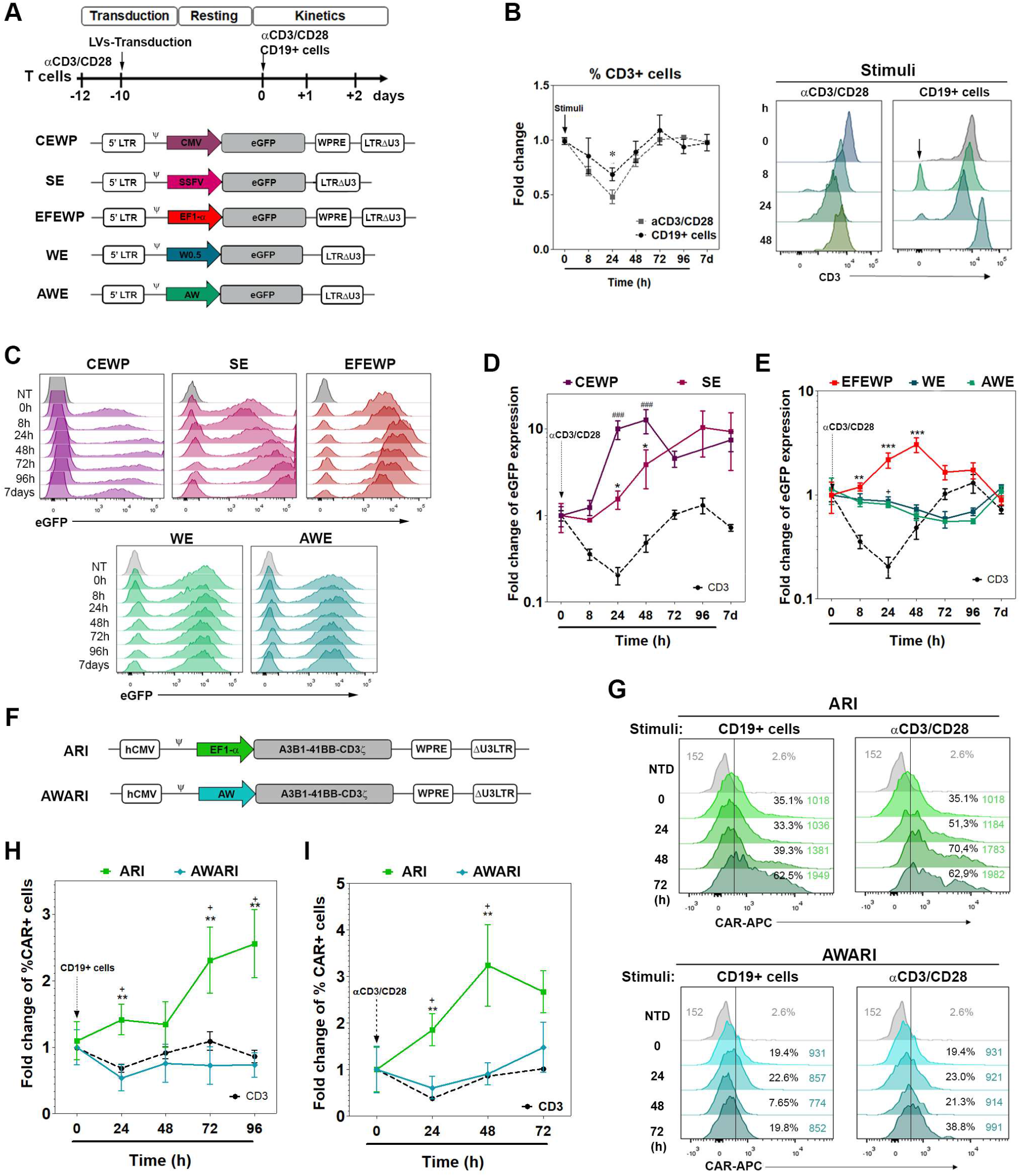
T cells transduced with *WAS* promoter-driven LVs mimic the TCR expression pattern. A) Top: Timeline of the experiments to study eGFP expression pattern after TCR/CAR stimulation. Bottom: diagrams of the different used eGFP-expressing LVs analyzed. B) Fold change of CD3 expression (% relative to 0h) in WT T cells stimulated with αCD3/αCD28 and αCD19CAR-T cells stimulated with CD19+ cells. Left: fold change in CD3 expression at different times post-stimulation. (Data are MeFI± SEM *, indicates p<0,05, 2-way-ANOVA, Bonferroni post-Test. n≥5). Right: Representative histograms showing CD3 expression on WT T cells (left) and aCD19CAR-T cells (right) at different times post-stimulation. C) Representative histograms of eGFP expression driven by strong-promoter driven (CEWP, SE and EFEWP-top panels) and *WAS-promoter* driven (WE, AWE - bottom panels) LVs at different times post-stimulation. D, E) Graphs showing fold change in eGFP (colored lines) and CD3 (black lines) expression along time after αCD3/αCD28 stimulation of T cells transduced with viral promoter-driven (CEWP, SEWP) (D) or eukaryotic-promoters driven (EFEWP, WE and AWE) LVs (E). Fold change is calculated by the following formula: ((MeFI of GFP+ population xh/ MeFI of non-transduced cells xh)/ MeFI of GFP+ population 0h/ MeFI of non-transduced cells 0h). 2-way-ANOVA, Bonferroni post-Test, compared with CD3 pattern: *, p<0.05; **, p<0.01; ***, p<0.001. +++ indicates p<0.001 for SEWP compared to CD3 expression. n=5. F) Diagrams of the EF1a promoter-driven (ARI) and *WAS*-promoter driven (AWARI) LVs expressing the ARI-0001 CAR (αCD19 A3B1-41BB-CD3ζ) G) Representative histograms of CAR expression levels on CAR-T cells generated with the ARI (top) or AWARI (bottom) LVs and measured at different time points after activation with CD19+ cells (left) or αCD3/αCD28 (right). H, I) Graphs showing fold change in CAR expression on ARI-CAR-T (green lines) and AWARI-CAR-T (blue lines) cells at different time points after activation with CD19+ cells (H) or aCD3/aCD28 (I). Data shows the percentage of CAR+ cells at different time points relative to 0 hours. Man-Whitney test, two tails, ** indicates p<0.01 compared to CD3 and + indicates p<0,05 compared to AWARI-CAR-T cells. n ≥ 5.

In order to analyse if the *WAS*-promoter-driven LVs mimic the TCR expression pattern, we first studied the CD3 expression levels (**Fig. 1B and Fig. S1a**) of primary human T cells after stimulation with αCD3/αCD28 nanomatrix and CD19+ cells (in CAR+ cells, **Fig 1B and Fig S1b**) at different time-points. In parallel, T cells transduced with the different eGFP-expressing LVs were stimulated and analysed by flow cytometry as described in **Fig. 1A**. As it has been reported previously^2^, CD3 expression in T cells decreased soon after stimulation reaching minimal expression at 24h/48h at both protein and mRNA levels and recover basal levels after 5-7 days (**Fig 1B, S1a**). Contrary to the CD3 expression pattern, all LVs harbouring constitutive-strong promoters (EF1α (EFEWP), SFFV (SE) and CMV (CEWP)), behaved in an opposite manner, increasing eGFP expression soon after T cell stimulation (**Fig. 1C**-top panels, **1D** and **1E**-orange line). Interestingly, *WAS*-promoter driven LVs partially mimicked the TCR expression kinetic, down-regulating eGFP expression upon stimulation that peak at 48h and recover basal levels after 7 days (**Fig. 1C**-bottom panels, **1E-**blue and green lines).

We next investigated whether CARs expressed by *WAS*-promoter driven LVs also followed a TCR-like kinetic. We constructed the AWARI LV (see M&M for details) based on the ARI-0001 LV backbone^21^, a CAR19-BBzz recently approved as ATMP in Spain, which uses the A1B6 clone as scFv (Fig. **1F**). We then generated ARI and AWARI CAR-T cells using the ARI and AWARI LVs and analysed their CAR expression kinetic upon activation with CD19+ cells **(Fig 1G**-left,**1H** and **S1B**-left) or αCD3/αCD28 (**Fig 1G**-right, **1I** and **S1B**-right). Interestingly, AWARI CAR-T cells showed a down-regulation of CAR levels after antigenic stimulation and a recovery after 48h (**Fig. 1H, 1I**, blue line; **Fig. 1G**, bottom panel), while ARI CAR-T cells showed a clear increment on CAR levels after T cell activation (**Fig. 1H, 1I**, green lines, **Fig. 1G**, top panel). A similar behaviour was also observed when using a third generation αCD19-CAR-28BBzz (**Fig S1C, S1D**). Altogether, our results showed that the AW-LVs can be used to generate CAR-T cells expressing the CAR in a TCR-like manner.

### TCR-like expression of CAR prevents exhaustion and control proinflammatory cytokines secretion upon antigen exposure *in vitro*

We next compared the behaviour of 2^nd^ generation ARI versus AWARI CAR-T cells. First, we measured basal levels of exhaustion markers (LAG-3, PD1, Tim3) (**Fig. 2A**) and tonic signalling (phospho-CD3z) (**Fig. 2B**) in a CD19-independent context. AWARI CAR-T cells expressed significant lower levels of LAG-3 and Tim3 (and also a trend for PD1) (**Fig. 2A**) and less phosphorylation of CD3z (**Figure 2B**). Both CAR-T cells efficiently killed CD19+ Nalm6 (B-ALL derived) and Namalwa (Hodgkin’s lymphoma derived), without significant differences (**Fig. 2C**). Interestingly, LAG-3 expression levels were significantly lower in AWARI compared to ARI CAR-T cells after killing Namalwa cells (**Fig. 2D**, right). In addition, AWARI CAR-T cells secreted lower levels of TNFα and IFNγ compared to ARI cells (**Fig. 2E**), indicating a more controlled proinflammatory activity. We also observed a tendency of increased stem memory T cells (CD45RA+CD62L+) population in AWARI versus ARI CAR-T cell (but not statically significant, **Figs. S2D, S2E**). Finally, similar observations were found when 3^rd^ generation CARs (CAR19-28BBzz) were expressed with the AW-LVs in terms of lower tonic signalling (**Fig S2A**) and milder IFNγ secretion (**Fig. S2B**).

**Figure 2.**
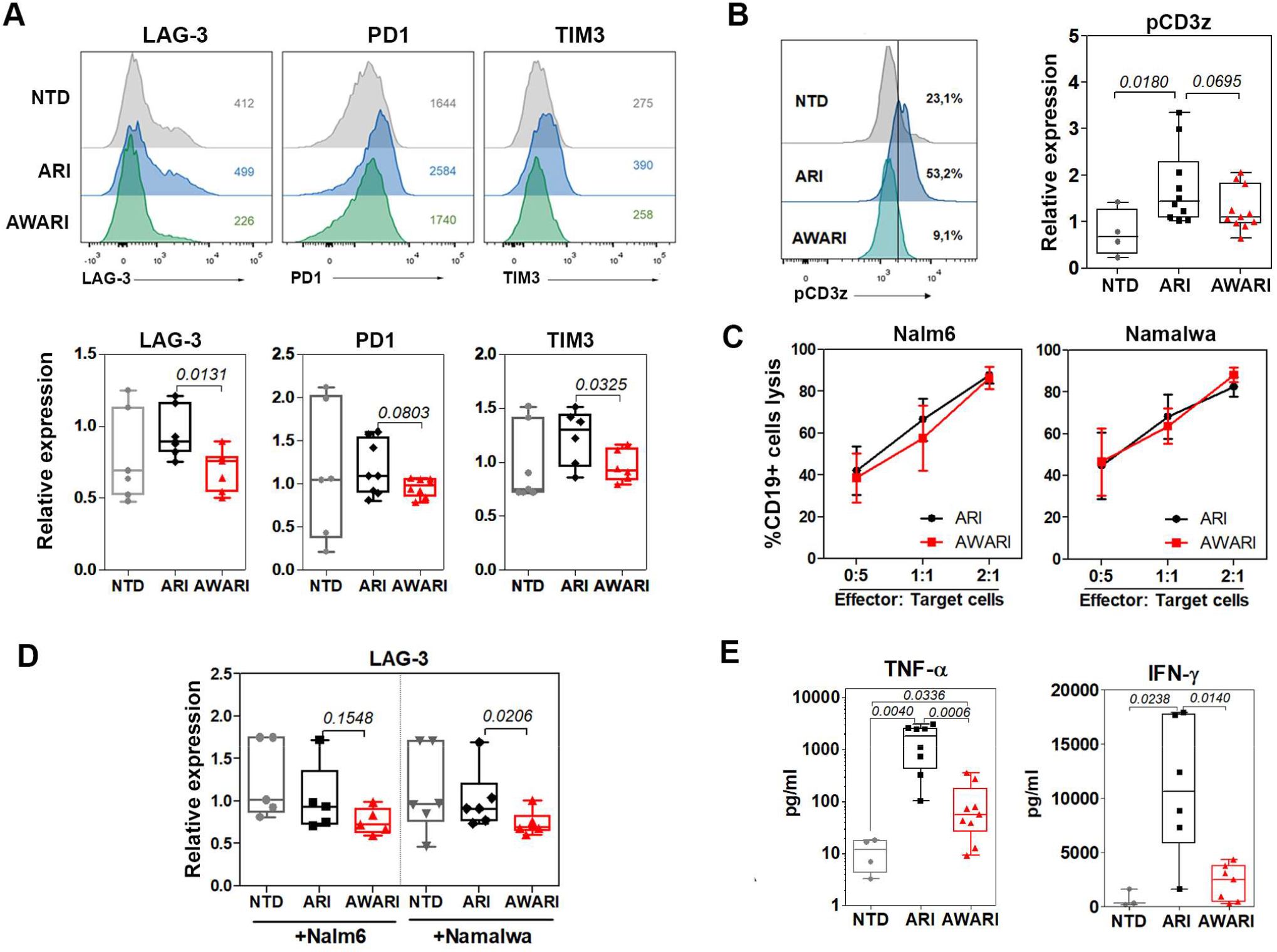
TCR-like expression improves CAR-T cells properties*in vitro*. A) Top: Representative histograms of LAG-3, PD1 and Tim3 expression of NTD (non-transduced, gray), ARI (blue) and AWARI (green) CAR-T cells determined 10 days after activation with αCD3/CD28. Bottom: Graphs showing expression levels of LAG-3, PD1 and Tim3 of ARI and AWARI CAR-T cells related to the expression in NTD cells. Data are MeFI ± SEM. One-tailed Man-Whitney test (4 independent-donors). B) Left: Representative histograms of intracellular phospho-CD3z expression after 10 days post-initial activation of NTD (gray), ARI (blue) and AWARI (green) CAR-T cells. Right: Graph showing pCD3z expression levels of each CAR-T cells related to NTD cells. Represented data is the ratio of pCD3z MeFI of ARI or AWARI CAR-T cells divided by the pCD3z MeFI of NTD cells. One-tailed Man-Whitney test (4 independent-donors, n≥4). C) Lysis of CD19+ cells (Nalm6 and Namalwa) at different ratios of co-culture with ARI and AWARI-CAR-T cells during 48h. Percentage of lysis is related to NTD-driven non-specific CD19+ lysis (n=5). D) Relative expression levels of LAG-3 on ARI and AWARI CAR-T cells 48h after co-culture with Nalm6 (left) or Namalwa (right) cells. Data represents LAG-3MeFI of ARI or AWARI CAR-T cells divided by the LAG-3 MeFI of NTD cells. One-tailed Man-Whitney test (n≥5). E) Levels of secreted TNFα (left) and IFN□ (right) produced by NTD, ARI and AWARI CAR-T cells after 24h of stimulation with Namalwa at ratio 1:1 and determined by ELISA (4 independent-donors, n≥4).

### AWARI CAR-T cells are as efficient as ARI CAR-T cells eradicating CD19+ tumour cells *in vivo* and maintain a more stem-like phenotype

*In vitro* cytotoxicity experiments only represent a direct-short term activity of CAR-T cells. We therefore performed a direct comparison of the anti-tumour activity of ARI versus AWARI CAR-T cells in a xenograft mice model of human lymphoma using two different CAR-T doses and a re-challenge after first lymphoma cells clearance (**Fig. 3A**, See M&M for details). Both CAR-T cells efficiently eliminated Namalwa tumour cells in treated mice with both doses (**Figs. 3B, 3C** and **Fig. S3A**). However, a small amount of Namalwa cells (less than <5%) were detected in liver of some ARI-mice and in brain of some AWARI-mice treated with the low dose (**Fig. 3C**, left panels). Importantly, CAR expression driven by the two vectors was maintained in all the organs after sacrifice (**Fig. 3D**), even with a second-challenge, when compared to those ex-vivo levels prior to infusion (**Fig. S3B**), indicating no promoter silencing.

**Figure 3.**
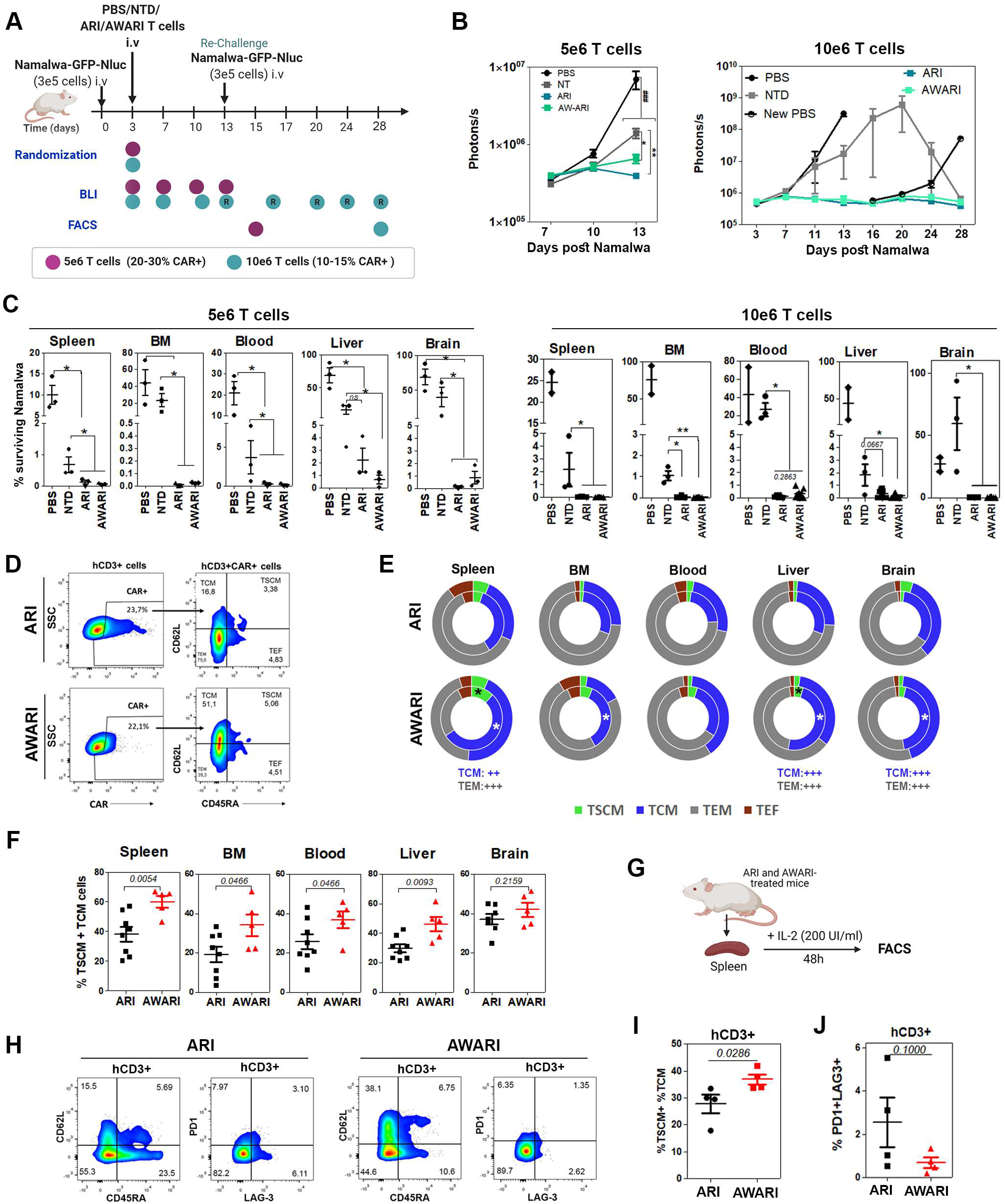
*In vivo* anti-tumor activity and phenotype of ARI and AWARI CAR-T cells. A) Experimental design of *in vivo* antitumor activity and phenotypic studies of 5e6 (purple dots) and a 10e6 (blue dots) of ARI and AWARI CAR-T cells. Re-challenge with new Namalwa is indicated with an arrow at day 16 in the 10e6 dose. Phenotypic analysis was performed after sacrifice at day 13 for 5e6 dose and day 28 for 10e6 dose. B) Tumor bioluminescence (BLI) progression with 5e6 (left) and 10e6 T cells doses (right) at different days post-Namalwa inoculation. C) Percentage of surviving Namalwa-GFP-Nluc cells in bone marrow (BM), spleen, blood, liver and blood determined by FACS after the sacrifice with the low (left panels) and high (right panels) doses. This value is analyzed in “singlets human cells” gate (see M&M). Two-tailed Mann-Whitney test, *, p<0.05; **, p<0.01, and ***, p<0.001. N(mice) of low dose: PBS=3, NTD=3, ARI=3, AWARI=3; and high dose: PBS=2, NTD=3, ARI=8, AWARI=5. D) Representative dot-plots of CAR expression and phenotype of CAR+ cells of gated hCD3 from the spleen of ARI and AWARI mice after sacrifice of the high-dose experiment. E) Graphical representation of proportions of T_N_/T_SCM_ (CD62L+CD45RA+) (green), T_CM_ (CD62L+CD45RA-) (blue), T_EM_ (CD62L-CD45RA-) (grey) and T_EF_ (CD62L-CD45RA+) (red) subpopulations in ARI (top) and AWARI (bottom) CAR+ cells in the indicated organs after sacrifice of challenged (outer circle) and re-challenged (inner circle) mice. *, p<0.05 when compared the T cells subsets of re-challenged ARI and AWARI treated mice. Mann-Whitney one-tail. ++ and +++ p<0.01 and p<0.001 respectively of 2-way ANOVA, Bonferroni post-Test of all the population of the total mice (ARI=8 (3+5R), AWARI=5 (2+3R). F) Percentage of T_N_/T_SCM_ and T_CM_ after sacrifice of ARI and AWARI hCD3+CAR+ cells (ARI=8, AWARI=5, Mann-Whitney one-tail. G) Spleen from treated mice were isolated and cells suspensions were cultured with 200 UI/ml of IL-2 in TexMACS medium and 2% P/S during 48h. H) Representative dot-plots of CD45RA/CD62L and PD1/LAG3 populations of ARI and AWARI mice. I) Percentage of hCD3+ T_N_/T_SCM_ and T_CM_after 48h of *in-vitro* culture. Two-tailed Mann-Whitney test (N=4). J) Percentage of PD1+LAG3+ cells. One-tailed Mann-Whitney test (N=4).

After showing the same therapeutic efficacy, we investigated potential differences in CAR-T cell phenotype after tumour clearance. Interestingly, AWARI-CAR-T cells showed a higher proportion of memory (T_CM:_ CD45RA-CD62L+) and stem memory (T_SCM_: CD45RA+CD62L+) (**Fig. 3D**) cells in spleen, bone marrow (BM), blood and liver when compared to ARI-treated mice (**Fig**.**3E, 3F**). Specially after re-challenge (inner circle, **Fig. 3E**), T_SCM_ population of AWARI-CAR-T cells was increased in spleen and liver and T_CM_ subset in BM and brain. In order to further characterize the potential differences, we cultured the cells from processed spleens during 48h *in vitro* with 200 UI/ml of IL-2 in TexMACs medium (**Fig. 3G**), and we confirmed an increment in T cell memory populations in AWARI-derived spleens (**Fig. 3H, 3I**) and a tendency to have lower expression of exhaustion markers (PD1+LAG3+) (**Fig. 3H, 3J**).

### Large-scale semi-automated manufacturing of AWARI CAR-T cells

Before translation of AW-LV to ATMP production for clinical applications, it is important to demonstrate their performance for large-scale automated manufacturing. We therefore used laboratory grade AWARI LVs to generate two batches of CAR-T cells (#01 and #02) in the CliniMACs Prodigy Bioreactor (Miltenyi Biotec) under GMP conditions (see M&M). Samples at days 7 and 10 for batch #01, and 6 and 9 days for batch #02 (**Fig. 4A**) were collected to study CAR expression (**Fig. 4B**), viability, expansion (**Fig 4C**), CD4/CD8 ratio (**Fig. 4D**), phenotype (**Fig. 4E**), and *in vitro* lytic activity (**Fig. 4F**). In both productions processes, cells were efficiently expanded, with more CD4+ than CD8+ CAR+ cells and prominent T_N_/_SCM_ (CCR7+ CD62L+) subset (44,85 ±0.07%) at day 7 or day 6, that were drastically reduced at final day of the production (16,55 ±13.93%), turning into T_CM_ cells but with minimal effector and effector memory populations (**Fig. 4E**). As described for other CAR-T cell products, lower expansion times rendered more potent AWARI CAR-T cells (**Fig. 4F**). All these results point out to the AWARI LVs as a valuable tool for a future clinical trial, although improvements in its lentiviral backbone could further increase transduction efficacy in order to lower costs and time of production.

**Figure 4.**
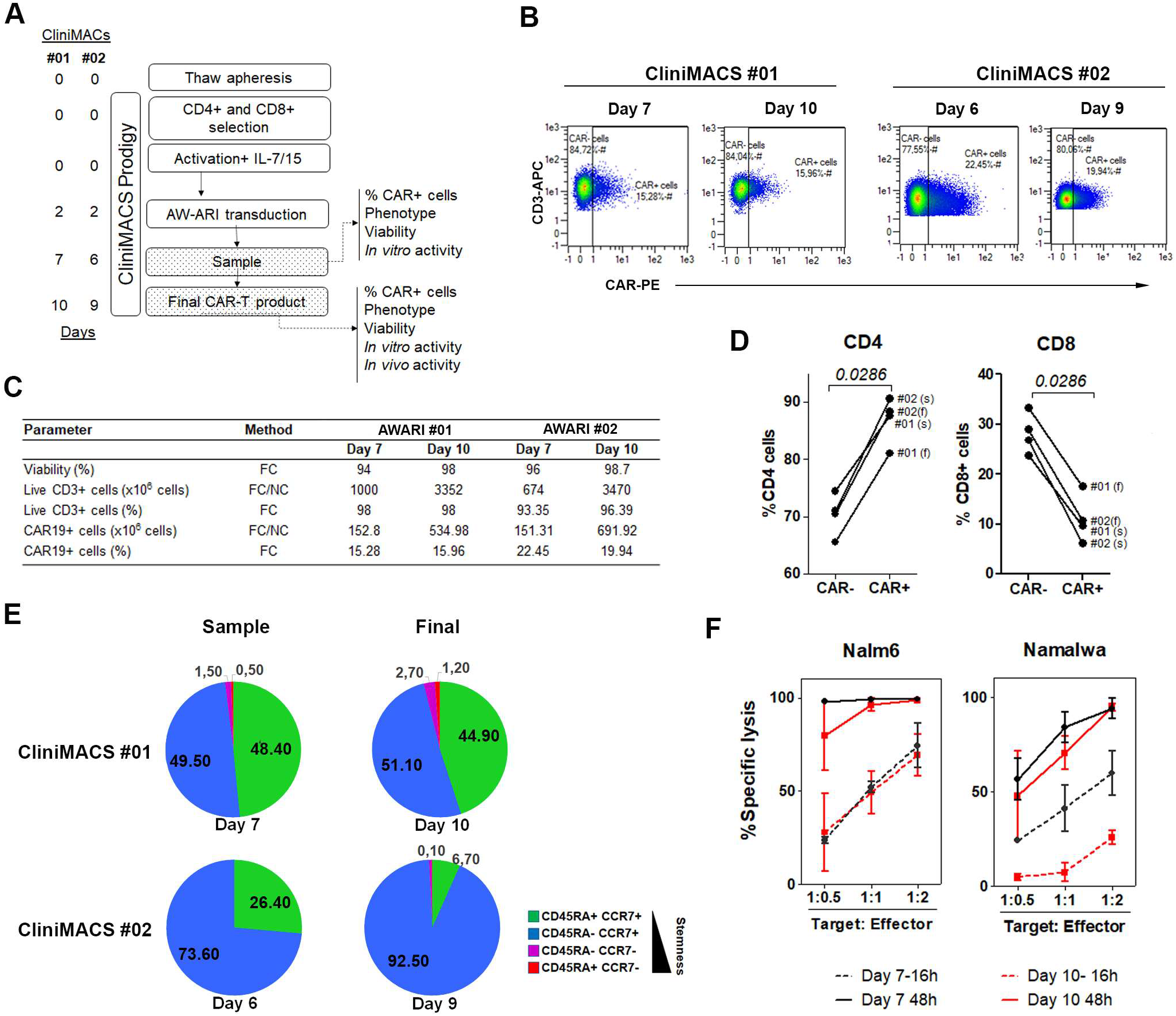
GMP-like large-scale production of AWARI CAR T cells. A) Time-line of the GMP-like batches #01 and #02 of AWARI CAR-T cells in the CliniMACS Prodigy. B) Dot-plots of CAR expression at days of sample (7/6) and final product (10/9) for both #01 (left) and #02 (right) batches. C) Data from viability, number of CD3, efficiency of CAR-T cell transduction and number of obtained CAR-T cells. FC= flow cytometry, NC= Neubauer chamber counting. D) Variation of percentage of CD4 and CD8+ cells in CAR- and CAR+ cells at sample (s) and final products (f). Two-tailed Mann-Whitney test. E) Graphical representation of the T cells populations at sample (days 6 or 7) and final products (day 9 or 10). T_N_/T_SCM_=Naïve/stem cell memory (CD45RA+CCR7+), T_CM_= central memory (CD45RA-CCR7+), T_EM_= effector memory (CD45RA-CCR7-) and T_EMRA_= effector memory RA+(CD45RA+CCR7-). F) *In vitro* specific lysis of CD19+ cells Nalm6 (left) and Namalwa (right) CD19+ cells by AWARI CAR-T cells harvested at sample day (black line) and final day (red line) for productions #01 and #02. CAR-T cells products were co-cultured at different ratios during 16h (dashed line) or 48h (continuous line).

### Large-scaled manufactured AWARI CAR-T cells products are highly efficient for tumour clearance and maintain good levels of T_N/SCM_ cells *in vivo*

Finally, we performed a detailed analysis of large-scale manufactured AWARI CAR-T cells in terms of their *in vivo* anti-tumour efficacy and their phenotype after isolation from treated mice (**Fig. 5**) for both #01 (blue dots) and #02 (green dots) CliniMACS batches. Anti-tumour activity of both AWARI CAR-T cells products were analysed in the Namalwa xenograft mice model as depicted in Fig 5A. Survival (**Fig. 5B**) and tumour progression (**Figs. 5C** and **5D**) were monitored up to 28-30 days post-treatment showing a strong anti-tumour activity of the AWARI CAR-T cells with 100% survival and undetectable tumour cells in treated mice (**Figs. 5C, 5D** and **Fig, S4A**). Control PBS and NTD-inoculated mice developed the disease at similar rates and the majority required to be sacrificed for compassionate reasons at days 13-16. At this time point, the stemness of human CAR-T cells of two AWARI-treated mice (**Fig. 5F** top panel, **S4C**) in #01 experiment was clearly maintained. New Namalwa cells were inoculated in AWARI-T cells-treated mice and also in new mice (“New PBS”). Re-challenged AWARI-mice remained tumour-free through the duration of the experiment as did the follow-up group. Similar results were observed in both productions (**Fig. 5C, 5D, S4A**). The percentage of CAR+ cells (**Fig 5E**) was maintained before *(ex-vivo)* and after *in vivo* elimination of Namalwa cells in the xenograf model with a slight increase in some of the organs (**Fig. S4b**). Importantly, the phenotypic analysis confirmed the presence of a high proportion of T_SCM_/T_CM_ populations (**Fig. 5E, 5F, S4D**) at the end point of rechallenged and non-rechallenged mice. However, a significant reduction of these populations was observed from day 13 to day 30 (**Fig. 5F top panel, S4C**). This data confirmed the *in vitro* (**Fig. S2**) and *in vivo* (**Fig. 3**) results showing that the expression of the ARI-CAR19-BBzz through a TCR-like promoter (AWARI-CAR-T cells) maintains the proportion of T_SCM_/T_CM_ CAR-T cells upon antigen encounter.

**Figure 5.**
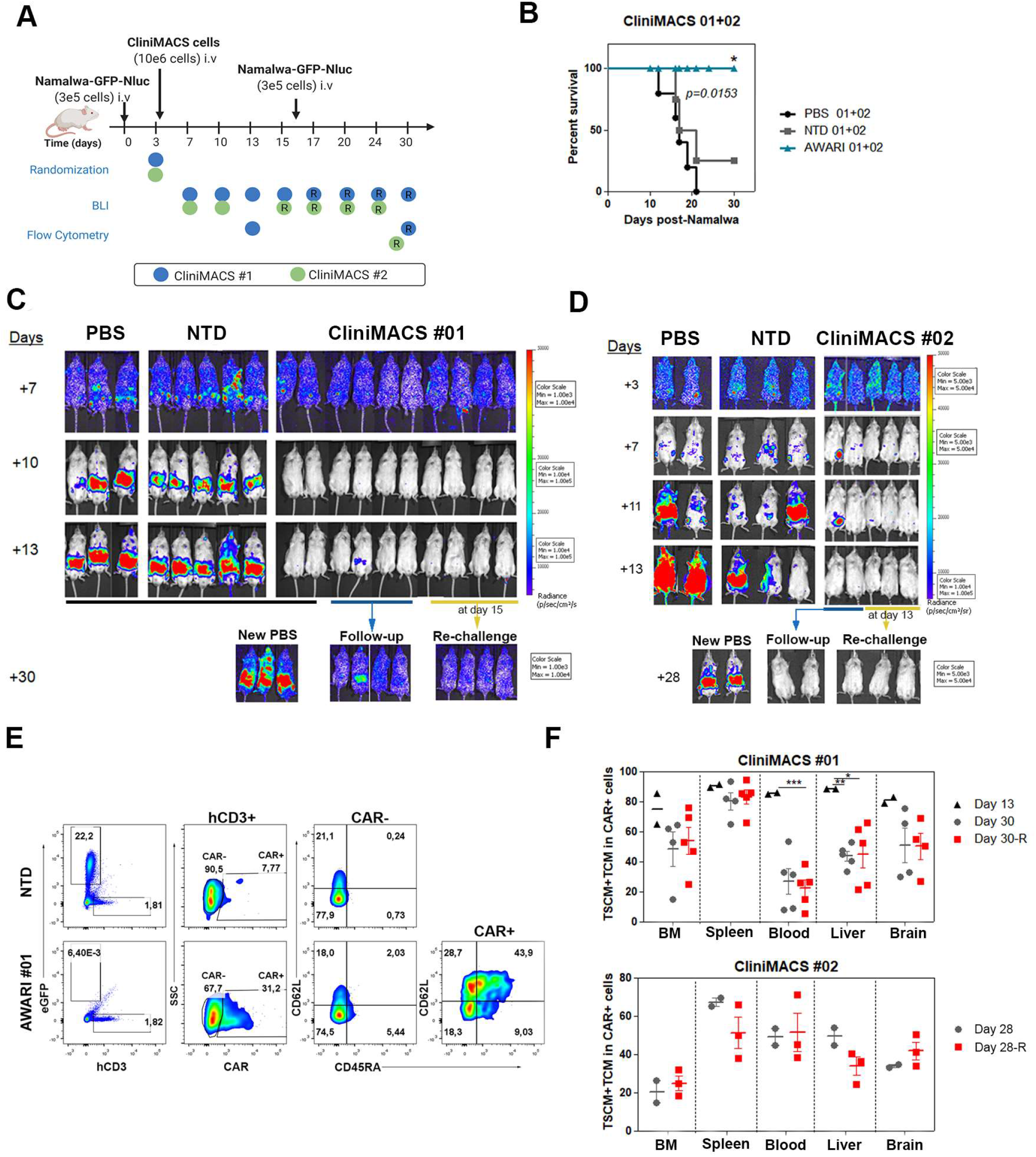
*In vivo* anti-tumor activity and phenotype of CliniMACS-produced AWARI-CAR-T cells after one or two Namalwa challenges. A) Experimental design and **t**ime-line of the two experiments with CliniMACS batches #01 and #02. R indicates re-challenge. B) Survival of PBS, NTD or AWARI-inoculated mice of #01 and #02 products. Log-Ranked test. C, D) Representative BLI images of tumor burden measured in #01 (C) and #02 (D) experiments at different days post Namalwa inoculation as indicated. In both studies, at day 15 (#01) or 13 (#02) respectively, new Namalwa cells were inoculated to AWARI treated mice (rechallenge, yellow line) and to control mice (‘New PBS’). E) Representative dot-plots of Namalwa/hCD3 presence, CAR+ cells of hCD3 and phenotype of CAR- and CAR+ of NTD cells and AWARI CliniMACS #01 from BM after sacrifice. F) Percentage of T_N_/_SCM_+T_CM_ population in hCD3+CAR+ cells analyzed in different organs from mice with one or two (rechallenge, R) inoculations of Namalwa tumor cells in #01 (left) and #02 (right) CliniMACS batches at final day. Only when CAR+ population was >1%, data was analyzed and included here. N (CliniMACS #01): PBS=3, NTD=5, AWARI=10 (2 sacrificed at day 13, 4 mice with one- and 4 mice with two-challenges. N (CliniMACS #02): PBS=2, NTD=3, AWARI =5 (2+3 re-challenged mice). 2-way-ANOVA, Bonferroni Post Test, *, indicates p<0.05, **, p<0.001, ***, p<0.0001.

## DISCUSSION

αCD19 CAR-T cells have achieved impressive therapeutic benefits in R/R B lineage lymphoid neoplasms, leading to the approval of Kymriah, Yescarta, Tecartus and Breyanzi and over 800 clinical trials around the globe. Most of these studies have used autologous T cells that are gene modified by retrovirus-based vectors to express the CAR through a strong promoter (often the EF1α or MSCV promoters). However, CAR-T therapy still has important limitations due to severe side effects and the lack of complete cures in 40-62% of the CD19+ lymphomas ^1^ and most solid tumours^22,23^. Improvements in CAR structure (scFV, hinge/spacer, costimulatory domains, bi-especific or multispecific CARs, etc) T cell composition or combinational therapies have already shown improved results^24^. New approaches to improve CAR-T efficacy and/or safety include the control of CAR activity and or other molecules^13^ and mechanism to avoid immune-reactions against the CAR-T cells^25^.

There are several evidences indicating that high levels and continuous expression of the CAR can be deleterious for the efficacy and safety of the CAR-T products^2-4^. The loss of efficacy comes in part from the early exhaustion and/or apoptosis of the CAR-T cells due to an excess of CAR signalling after antigen encounter^26^. Strong and unregulated CAR expression also increase tonic signalling in the absence of target antigen^4^ promoting exhaustion^3, 5^. On the other side, safety issues due to CAR overexpression relates to the increased antigen-independent tonic signalling and to over-stimulation of CAR-T cells that generate CRS. Using genome editing technologies, Eyquem et al. demonstrated that TCR-like expression of CARs resulted in lower tonic signalling, reduced CAR-T exhaustion and increased proportion of TSM/TN CAR-T cells, resulting in improved anti-tumour activity^2^. These authors also showed that low expression was not enough to achieve optimal CAR-T products, concluding that a down-regulation of the CAR upon antigen encounter was required for optimal CAR-T performance.

In this manuscript we investigated whether LVs can be used to obtain a TCR-like expression kinetics that could also result in an improved CAR-T cell product. Our search for human promoters that could mimic the TCR expression pattern pointed to the TRAC locus, however, the TRAC promoter in mature human T cells is not well defined. We therefore focus on the *WAS* gene promoter because WASP protein is only expressed in hematopoietic cells, is involved in the formation of the immunological synapse and acts as an adaptor of TCR signals^14,15^. We have previously developed different *WAS*-gene based promoters and identified the AW promoter, harbouring fragments from the alternative and proximal promoter, as the best option to express transgenes in hematopoietic cells^8,11,16^. Here we showed that, indeed, *WAS*-promoter driven LVs (AW-LVs) partially mimicked the TCR expression kinetic, with a small downregulation upon stimulation and recovering basal levels in 5-7 days.

Based on the ability of AW-LVs to mimic TCR expression pattern on T cells, we investigated whether 4-1BB-CAR-T cells generated with this “physiological” LVs have a better behaviour compared to that generated with LVs harbouring strong or viral promoters. Although in terms of *in vitro* or *in vivo* anti-tumour activity we could not detect significant differences with EF1α-driven CAR-T cells, AW-CAR-T cells exhibited lower tonic signalling and a better phenotype after efficient killing of CD19+ cells, with reduced secretion of pro-inflammatory cytokines. These data is largely in agreement with the observations of Eyquem et al. using genome editing technologies to express the CAR through the TRAC locus^2^. However, these authors also showed improved *in vivo* anti-tumour activity^2^. The differences in the tumour model (Nalm6 versus Namalwa) as well as in the CAR signalling domain (CD28 versus 4-1BB) and scFv (FMC63 versus A3B1) could explain the observed differences. Indeed, the potential deleterious effects of CAR over-expression and tonic signalling are highly influenced by the CAR configuration^2, 4^. For instance, replacing CD28 with the 4-1BB costimulatory domain reversed exhaustion in CAR T cells and improved its persistence and therapeutic efficacy^27^. The good behaviour of EF1α-driven 4-1BB-CARs T cells and the experimental limitations derived by the graft versus host disease (GVHD) caused by the administration of the CAR-T cells can explain the absence of increased therapeutic efficacy in our animal models.

In spite of the good behaviour of 4-1BB-CARs, Gomes-Silva et al demonstrated that very high expression levels of 4-1BB-CARs also lead to apoptosis and limit CAR-T cells expansion^4^. The same authors also showed that lowering it expression levels by using EF1α-driven LVs instead of LTR-driven □-retroviral vector, improved phenotype, expansion potential and therapeutic efficacy. These studies showed that EF1α-driven LVs already generate a good CAR product for some 4-1BB-CARs. Here we show that TCR-like expression can further improve 4-1BB-CAR-T products, in particular for the ARI-0001, by maintaining a better phenotype and controlling tonic signalling and pro-inflammatory cytokine secretion.

An important aspect to consider in CAR-T cells is the potential side effects due to overstimulation and/or inadequate stimulation. Severe CRS (grades 3 or 4) is an important hallmark in CD19+ that can compromise efficacy and lead to life-threatening conditions^28,29^. Upon tumour cell encounter, CAR-T cells release different pro-inflammatory cytokines such as TNFα, IFN□, IL-1 and GM-CSF that leads to macrophage recruitment and activation causing a dangerous cytokine storm involving IL-6 and GM-CSF^30, 31^. It has also been demonstrated that activation of macrophages through CD40/CD40L, CD69 and LAG-3 also contributes to massive activation^29^. Although CRS can be managed with corticosteroids and anti-IL6R (Tocilizumab), these interventions also block T cell activation compromising CAR-T efficacy ^29^. In this manuscript, we have shown that CAR-T cells generated with AW-LVs exhibit a milder secretion of TNFα and IFN□ after tumour cells encounter. In addition, we noted that LAG-3 expression was reduced after Namalwa interaction which suggests that AW-CAR-T cells could lower the risk of CRS.

We finally investigated the feasibility of generating a viable CAR-T cell product for clinical translation. Here, we have demonstrated that it is feasible to generate TCR-like CAR-T cells in a CliniMACs Prodigy, with high cell viability, predominant stem and CM populations and potent *in vitro* and *in vivo* anti-tumour activity. In contrast and consistent with our comparative results, ARI-0001 generated more memory and effector memory population^19^, highlighting another potential advantage of AW-CAR-T cells during the manufacturing process. On the other hand, the efficacy of transduction was lower compared to ARI (∼ 18.4% vs ∼30.6%), suggesting that a future optimization of the AWARI LV backbone could further improve the manufacturing process of AW-CAR-T cells.

Taken all together, we propose the use of AW-LVs as an alternative platform for the manufacturing of CAR-T cells in order to provide better efficacies and/or safety of the final products. The final benefits of expressing different CARs through the AW promoter will need to be determined in each case.

## Conclusions

In this work we showed that LVs expressing the transgene through the *WAS* gene proximal promoter mimic very closely the TCR expression pattern kinetic in T cells. 4-1BB-CAR-T cells generated with *WAS-*promoter-driven LVs (AWARI CAR-T) exhibited a higher proportion of Naïve/Stem Cell Memory T cells, less exhausted phenotype, lower tonic signalling and reduced secretion of pro-inflammatory cytokines compared to EF1-driven ARI CAR-T. Finally, AWARI CAR-T cells were efficiently manufactured in large-scale GMP-like conditions. Altogether, our data showed that the AW-LVs can be used to generate improved, physiological 4-1BB-CAR-T cell products and propose these LVs as efficient tools for the generation of clinical-grade TCR-like CAR-T cells.

## Data Availability

All data relevant to the study are included in the article or uploaded as online supplemental information.

## DECLARATIONS

### Ethics approval and consent to participate

All donors from the Haematology Unit of Hospital Reina Sofía, Córdoba (Spain) gave their written informed consent and the study was performed according to the established guidelines of in the approved by the Regional Government Junta de Andalucía-Consejeria de Salud (date of approval: 22/02/2019, signed by D^a^ Cristina Lucía Dávila Fajardo as secretary of the Research Ethics Committee), that comply with the requirements regarding quality and safety for donation, obtaining, storage, distribution, and preservation of human cells and tissues under the Spanish specific regulation (RD 9/2014) and International Conference of Good Clinical Practice. All the experiments involving animals were performed according to a protocol approved by the Institutional Animal Care and Use Committee of the University of Granada (13/12/2016/181) were in accordance with the European Convention for the Protection of Vertebrate Animals used for Experimental and Other Scientific Purposes (CETS # 123) and the specific Spanish law (R.D. 53/2013). Subjects were randomly assigned to receive the different treatments and sample size estimation was calculated based on Mayer et al^20^.

### Consent for publication

All the authors consent the publication of this manuscript.

### Availability of data and material

All data relevant to the study are included in the article or uploaded as online supplemental information. Figures were created with Biorender.com.

### Authors’ contributions

MTM and NMP designed and performed experiments, analysed data and wrote the manuscript. PLJL performed and analysed experiments and critical review of the manuscript. PM contributed to animal experiments. MCG and KP contributed to T cells isolation and revised the manuscript. RJM and SN designed and produced CliniMACs Prodigy large scale CAR-T cells production process. RJM, SN and MDC performed the phenotypic characterization of CliniMACs-derived products. SSH and AAG contributed to AWARI LVs production. MC and MJ provided ARI-0001 LV and critical review. CM provided material and critical review. KB provided funding and critical review. CH provided funding and critical review of the manuscript. FM conceals the project, designed experiments, analysed data, provides the main funding and wrote the manuscript.

## Acknowledgements

We thank Ana Fernández-Ibáñez for her support with the IVIS Spectrum Analyzer, Nieves Álvarez for her help of ELISA assays. We also thank GENYO’s supporting units.

## Physiological (TCR-like) regulated lentiviral vectors for the generation of improved CAR-T cells

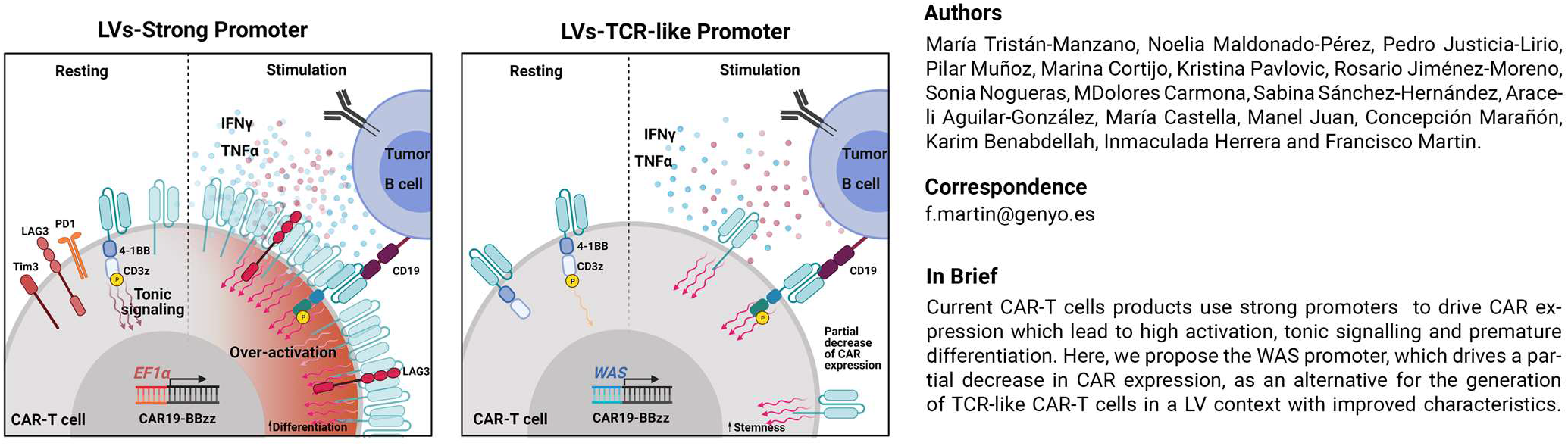

## List of abbreviations

ATMP: advanced therapy medical product
B-ALL: B-acute lymphoblastic leukaemia
CAR: Chimeric Antigen Receptor
CMV: Cytomegalovirus
eGFP: enhanced green fluorescent protein
EF1α: Elongation Factor 1-α
EMA: European Medicine Agency
FDA: Food and Drug administration
LVs: Lentiviral vectors
LTR: long-terminal repeats
MSCV: murine stem cells virus
NTD: non-transduced cells
R/R: relapsed/ refractory
ScFv: Single chain variable fragment
SFFV: Spleen focus forming virus
TCR: T cell receptor
TCM: T central memory
TEF: T effectors
TEM: T effectors memory
TN/SCM: T naive/stem cell memory
TRAC: T cell receptor (TCR) constant alpha locus
WAS: Wiskott-Aldrich syndrome

