## Supplementary figures and images for "Physiological (TCR-like) regulated lentiviral vectors for the generation of improved CAR-T cells"

### Figure S1

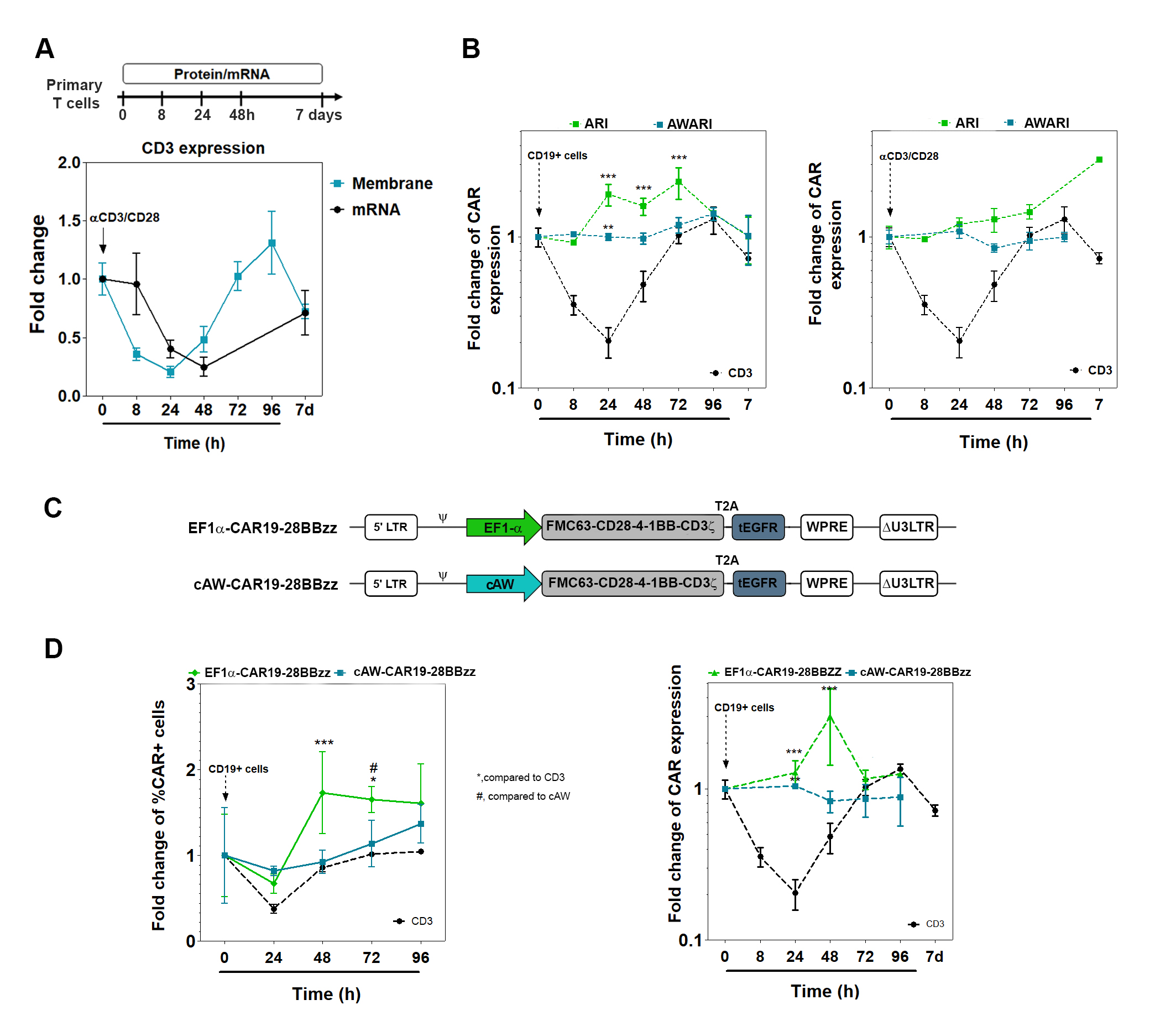

### Figure S2

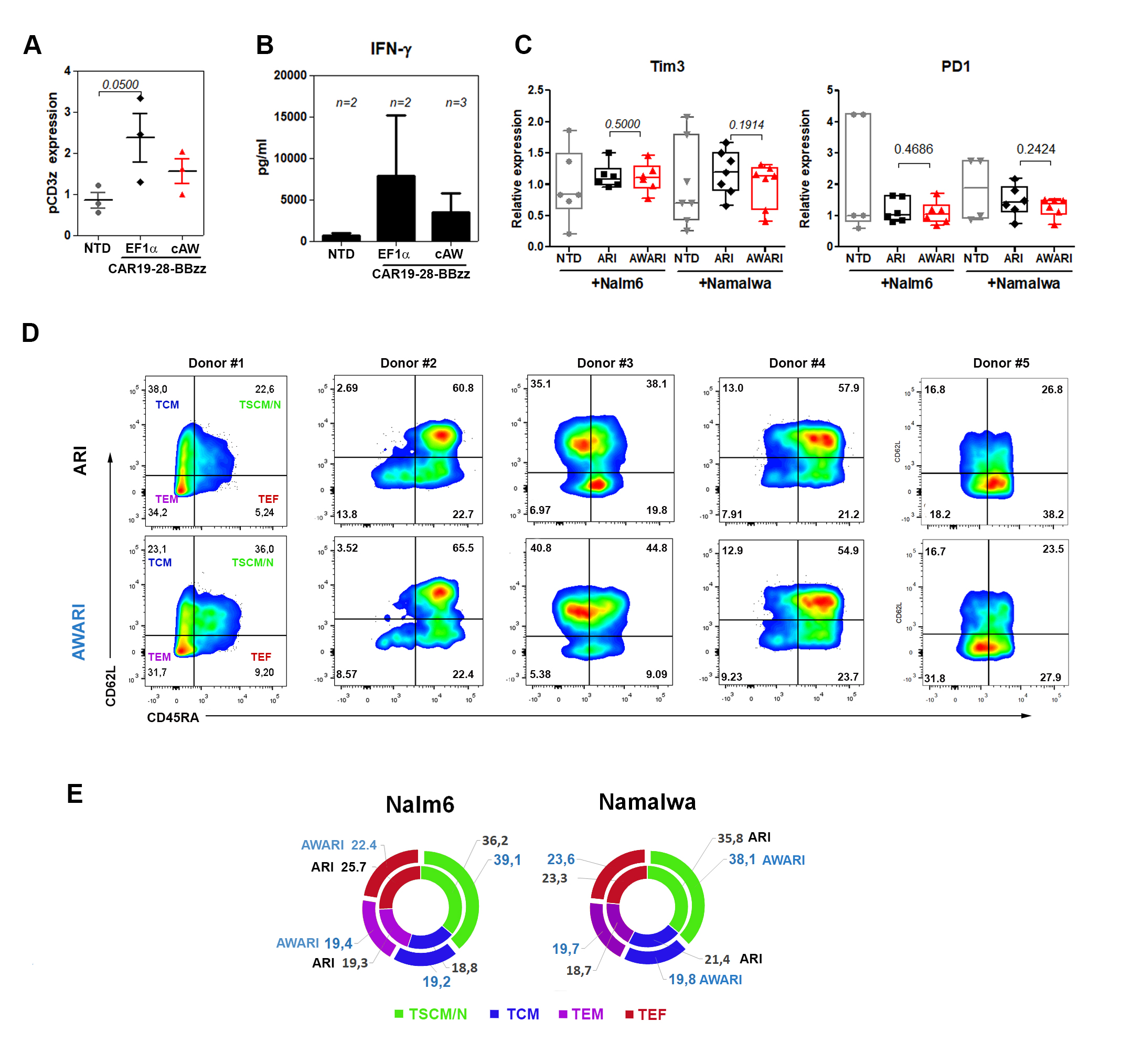

### Figure S3

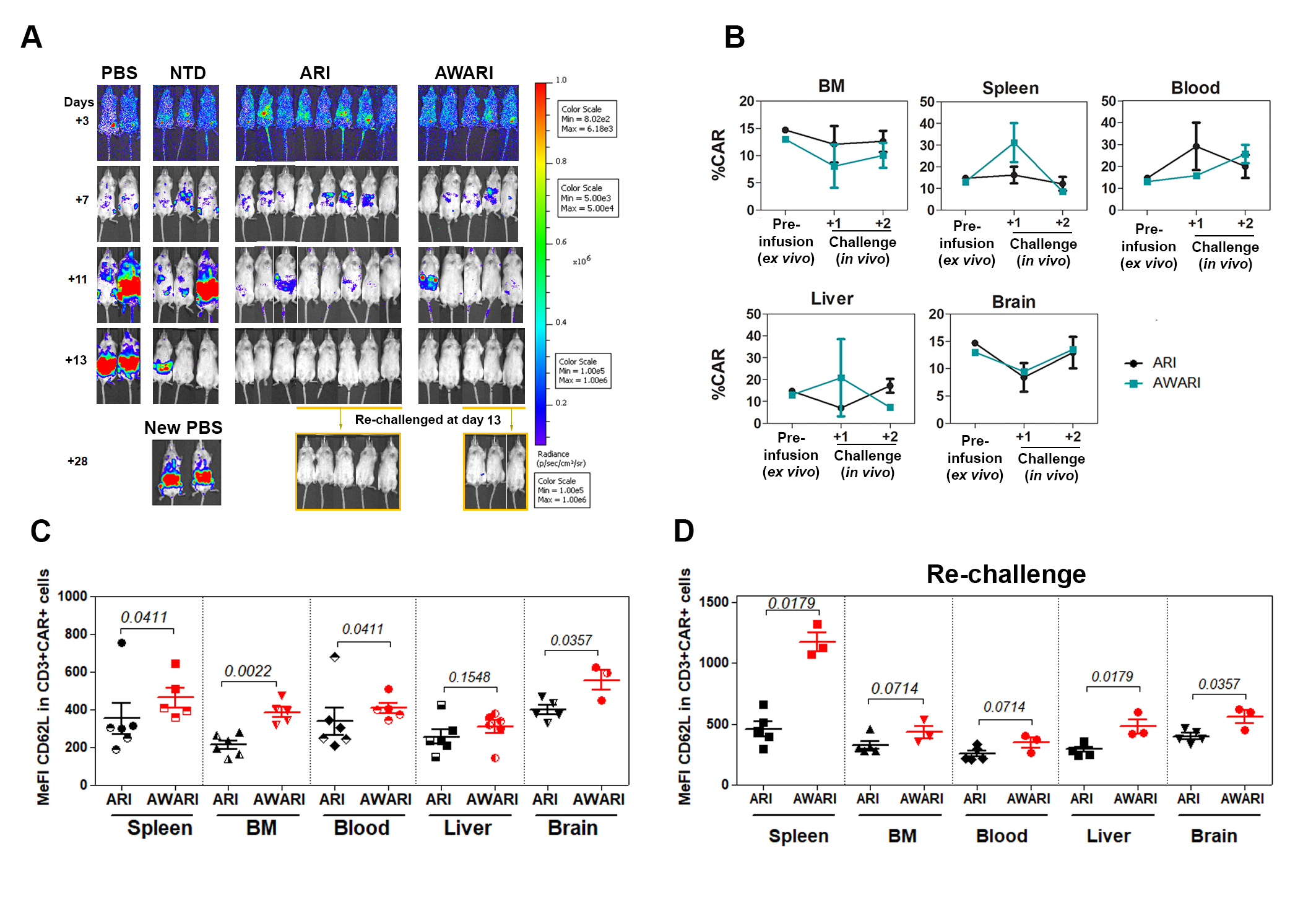

### Figure S4

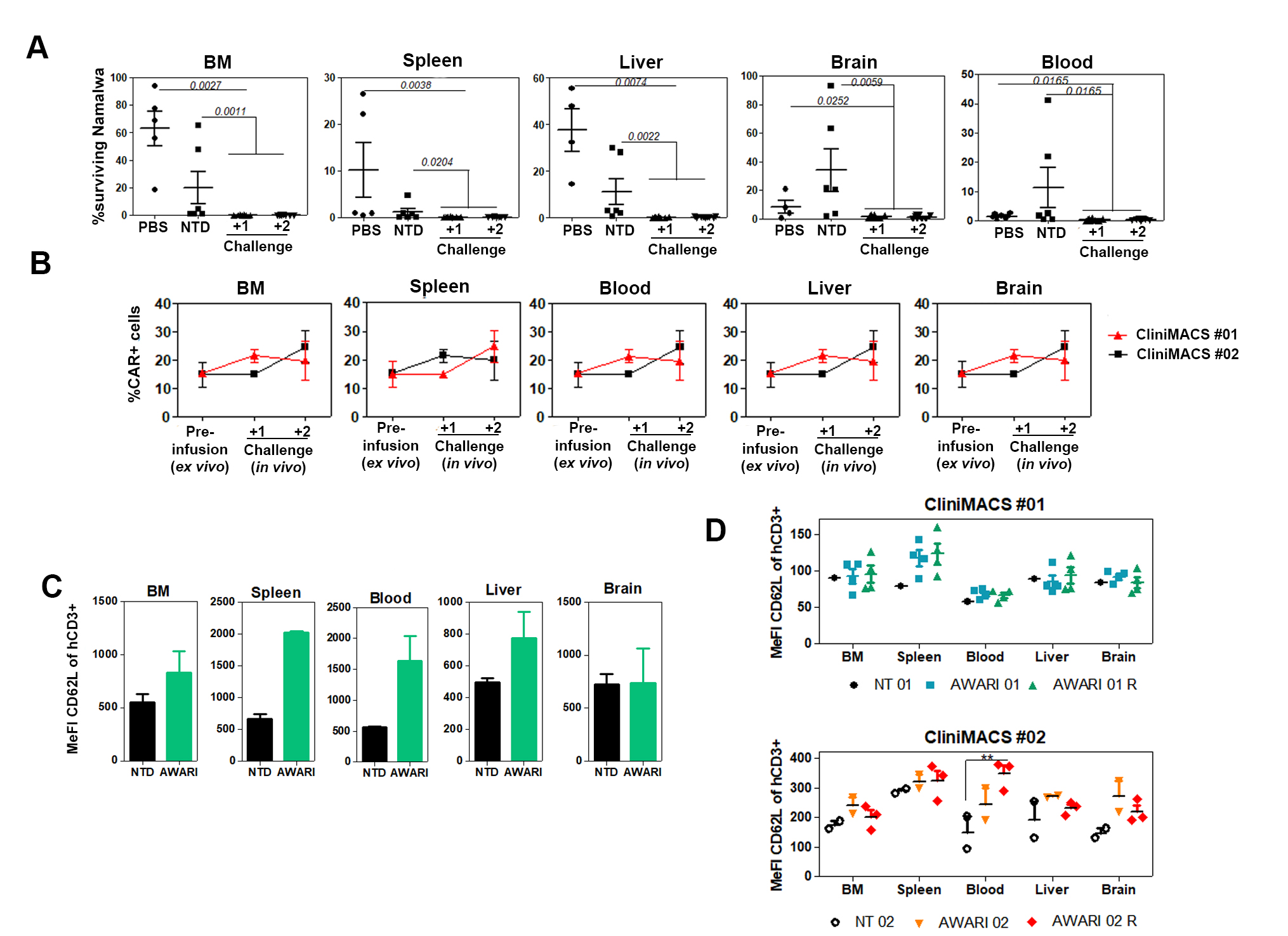
