## Supplementary text for "Physiological (TCR-like) regulated lentiviral vectors for the generation of improved CAR-T cells"

**SUPPLEMENTARY FIGURES**

**Supplementary Figure 1.** A) Fold change of CD3 protein (blue) and mRNA (black) expression (% relative to 0h) in WT T cells stimulated with αCD3/CD28 (n≥5). B) Fold change of CAR expression in ARI (green) and AWARI (blue) CAR-T cells stimulated with CD19+ cells (left graph) and αCD3/CD28 (right graph). Fold change of CD3 expression is shown in black dashed lines in both graphs as reference (n ≥5). Expression was calculated as CAR MeFI of CAR+ population/ CAR MeFI of non-transduced T cells. Fold change indicates the expression at different times related to those of 0h. C) Diagrams of 2^rd^ generation LVs expressing 3^rd^ generation αCD19 (FMC63) CARs (CAR19-28BBzz) through the EF1α (top) and *WAS* (bottom) promoters. In both cases, a truncated EGFR is expressed through the T2A cleavage peptide. D) Fold change of % CAR+ cells (left) and CAR expression levels (right) driven by EF1α-driven (green lines) and *WAS*-promoter driven LVs expressing CAR19-28BBzz after CD19+ stimulation (n=5). ANOVA test, Bonferroni Post-test. *, compared to CD3; #, compared to *WAS* promoter.

**Supplementary Figure 2.** A) Phospho-CD3z expression of non-transduced (NTD) T cells and CAR19-28BBzz-T cells generated with EF1α- and *WAS*-promoter driven LVs. Data shows pCD3z MeFI relative to those of NTD. One-tailed Man-Whitney test (three independent donors). B) IFN-ɣ secretion from EF1α- and *WAS-*of CAR-T-28BBzzcells after incubation with CD19+ target cells for 24h at ratio 1:1. C) Relative expression of Tim3 (left) and PD1 (right) driven by 2^nd^ generation ARI and AWARI CAR-T cells after being co-cultured with Nalm6 or Namalwa during 48h in the presence of HL-60 cells. One-tailed Man-Whitney T test, no significative. D) Dot-plots of CD45RA/ CD62L of ARI and AWARI CAR-T cells from five different donors after 48h in culture with Namalwa cells at ratio 1:1. E) Graphical representation of the different T cells subsets of ARI (black) and AWARI (blue) after 48h of Nalm6 and Namalwa (n=5) exposition. ANOVA-T test, no significant.

**Supplementary Figure 3.** A) BLI images of PBS, NTD, ARI and AWARI treated mice up to 28 days. Re-challenged mice are highlighted in yellow. B) Percentage of CAR+ cells of the hCD3+ population in ARI and AWARI-CAR-T cells pre-infusion *(ex vivo)* and 28 days after their inoculation into mice with one (+1) or two (+2) Namalwa challenges *in vivo*. Data for bone marrow (BM), spleen, blood, liver and brain are shown. C) CD62L MeFI of ARI and AWARI hCD3+CAR+ cells with only one *in vivo* challenge of Namalwa cells. Data from the two experiments using the low and high doses (5e6 and 10e6) from figure 3 are compiled here. Half-filled dots indicate data from 5e6 dose and filled dots from the 10e6 dose. One-tailed Mann-Whitney. N(5e6 dose): ARI=3, AWARI=3. N(10e6 dose): ARI=3, AWARI=2. Only when CAR+ population was >1%, data was analyzed and included here.

**Supplementary Figure 4.** A) Percentage of surviving Namalwa cells of both #01 and #02 CliniMACS experiments with one or two *in vivo* challenges with Namalwa-GFP-Nluc. Data were analyzed by FACS at final point. B) Percentage of CAR+ cells inside the hCD3+ population in AWARI-CAR-T cells from the two CliniMACS productions analyzed *ex vivo* (pre-infusion) and 28 days after their inoculation into mice with one (+1) or two (+2) Namalwa challenges. Data from bone marrow (BM), spleen, blood, liver and brain are shown. C) CD62L MeFI of NT and AWARI CAR-Ts from mice inoculated with #01 production at day 13 of the first Namalwa infusion. N (NTD)=2 and N(AWARI)=2. D) CD62L MeFI of hCD3+ from different analyzed organs of NT and AWARI mice with one or two (re-challenged, R) tumor inoculations of #01 (top) and #02 (bottom) at final day of the experiments.

**Supplemental material**

**RNA extraction and quantitative PCR**

Total RNA from T cells was extracted using Trizol (Ambion) and RNA samples were converted into cDNA using the high-capacity cDNA reverse transcription kit (ThermoFisher), complemented with RNase inhibitor (ThermoFisher). Q-PCR was performed in a Stratagene MX3005P system, using the Kappa SYBR FAST pPCR Master Mix (Kappa Biosystems) and primers against CD3: CD3-Fw 5´- AAGATGAAGTGGAAGGCG-3´ and CD3-Rv 5´-CTCAGGAACAAGGCAGTG-3´. GAPDH as housekeeping gene: GAPDH-Fw 5´-ATGGGGAAGGTGAAGGTCG-3´ and GAPDH-Rv 5´-GGGGTCATTGATGGCAACAATA-3´. Relative changes in gene expression were analysed using the 2^ΔΔCt method.
